# Systematic review of distribution models for *Amblyomma* ticks and Rickettsial group pathogens

**DOI:** 10.1101/2020.04.07.20057083

**Authors:** Catherine A. Lippi, Holly D. Gaff, Alexis L. White, Sadie J. Ryan

## Abstract

The rising prevalence of tick-borne diseases in humans in recent decades has called attention to the need for more information on geographic risk for public health planning. Species distribution models (SDMs) are an increasingly utilized method of constructing potential geographic ranges.There are many knowledge gaps in our understanding of risk of exposure to tick-borne pathogens, particularly for those in the rickettsial group. Here, we conducted a systematic review of the SDM literature for rickettsial pathogens and tick vectors in the genus *Amblyomma*. Of the 174 reviewed papers, only 24 studies used SDMs to estimate the potential extent of vector and/or pathogen ranges. The majority of studies (79%) estimated only tick distributions using vector presence as a proxy for pathogen exposure. Studies were conducted at different scales and across multiple continents. Few studies undertook original data collection, and SDMs were mostly built with presence-only datasets from public database or surveillance sources. While we identify agap in knowledge, this may simply reflect a lag in new data acquisition and a thorough understanding of the tick-pathogen ecology involved.

## Introduction

Tick-borne diseases are a global threat to public health, posing risks to both humans and domesticated animals. In recent years there have been documented increases in tick-borne diseases both in the United States and around the world. Much of this burden can be attributed to Lyme disease in the United States, Europe, and northern Asia. However, in the past 20 years,identification of previously unrecognized pathogens has revealed a great diversity in tick-borne viruses and bacteria (Paddock et al., 2016). Increases in tick-borne pathogen transmission and case detection have garnered a great deal of attention, triggering greater funding, resources, and agency responses (CDC, 2018; Couzin-Frankel, 2019). Nevertheless, the expanding burden of tick-borne disease has also highlighted crucial gaps in knowledge, particularly with regards to geographic risk mapping, an area of great interest to public health agencies. This is particularly evident in the case of rickettsial pathogens, comprising the ehrlichiosis, anaplasmosis, and spotted fever rickettsioses, which compared to Lyme disease remain understudied. Rickettsial pathogens of medical importance may be encountered worldwide, and ticks from the *Amblyomma* genus are competent vectors for many of these pathogens (Levin et al., 2018).Although numbers of documented cases have been increasing in recent years, the true extent of geographic risk for rickettsial pathogens is challenging to delineate due to a lack of consistent,long-term, and widespread surveillance data, and regionally low case detection.

Species distributions models (SDMs), also commonly referred to as ecological niche models (ENMs), are becoming routinely used in vector-borne disease systems to model the potential geographic distribution of risk (e.g. Baak-Baak et al., 2017; Carvalho et al., 2015; Lippi et al., 2019; Peterson et al., 2002; Thomas and Beierkuhnlein, 2013). Broadly, this is accomplished by correlating locations where a species of interest is known to occur with the underlying environmental characteristics (e.g. climate, elevation, land cover). The resulting model can then be projected to unsampled areas on the landscape, providing a spatial prediction of areas that are ecologically suitable for species presence. In addition to predicting contemporary species distributions, SDMs are also employed to estimate the extent of potentially suitable habitat for invasive species, and potential shifts in geographic distributions due to climate change (Lippi et al., 2019). There are many methodological approaches to estimating species distributions, and some of the more commonly encountered approaches include Maximum Entropy (MaxEnt), Generalized Additive Models (GAM), Boosted Regression Trees(BRT), and Random Forests (RF) (Elith et al., 2008; Elith and Leathwick, 2009; Evans et al.,2011; Phillips et al., 2006). Although SDMs are a commonly used tool in estimating species ranges, the diversity in modeling approaches and applications makes it challenging to compare results across models.

For vector-borne disease SDMs, records of vector and/or pathogen presence (either the vector, the pathogen, the vector and pathogen, or even simply human case data) are often used as proxies for risk of exposure, and therefore transmission. Species distribution models have been used in a risk mapping capacity for many vector-borne disease systems, spanning a range of pathogens vectored by arthropods including mosquitoes, gnats, phlebotomine flies, fleas,triatomine bugs, and ticks (Crkvencic and Šlapeta, 2019). This framework is particularly useful in determining species limits for vectored transmission owing to the very close relationships between ectotherm life histories, pathogen replication, and environmental drivers such a temperature. Underlying distributions of reservoir hosts, another requisite component of zoonotic transmission cycles, are also determined by land cover and environmental conditions.

This work provides a comprehensive review of the published, peer-reviewed literature of studies that estimated species distribution, or ecological niche, of *Amblyomma* ticks, the rickettsial pathogens they vector, or their combined distributions. Following Preferred Reporting Items for Systematic Reviews and Meta-analyses (PRISMA) guidelines, we identified and compiled studies that used occurrence records and environmental predictors to estimate the geographic range of target organisms (Moher et al., 2009). Additionally, we provide a synthesis of current knowledge in the field, identifying the range of regions, spatial scales, and environmental determinants used to define risk in these systems. This work serves as a baseline for identifying knowledge gaps and guiding new studies of geographic risk mapping in understudied tick-borne disease systems.

## Materials and Methods

Literature searches were conducted following the guidelines in the PRISMA Statement, a checklist and flow diagram to ensure transparency and reproducibility in systematic reviews and meta-analyses (Liberati et al., 2009; Moher et al., 2009). Initial searches for peer-reviewed studies were conducted through September 2019. Five online databases were searched including Web of Science (Web of Science Core Collection, MEDLINE, BIOSIS Citation Index,Zoological Record) and Google Scholar. Searches were performed with combinations of key terms including “Amblyomma”, “Rickettsia*”, “niche model”, “ecological niche model”, and “species distribution model”. No restrictions were placed on geographic region of study or date of publication. Additional novel records for screening were identified via literature cited sections in records identified via database searches.

Duplicate records from the initial database searches were removed, and the remaining abstracts were screened for relevance. Records were excluded in this first screening based on publication type (i.e. literature reviews, opinion pieces, and synthesis papers were excluded), methodology (i.e. studies that did not examine geographic distributions or risk were excluded), and target organisms (i.e. vectors and diseases other than the *Amblyomma* spp. and rickettsial pathogens (*Ehrlichia* spp. and *Rickettsia* spp.) were excluded). *Anaplasma* spp. were excluded from our analysis as these are primarily transmitted by *Ixodes* ticks.

The remaining articles were then assessed in full for eligibility, where information on vectors, pathogens, modeling methods, geographic region, geographic scale of study, time period of study, data sources, and major findings were extracted from the text. Full-text articles in this final screening were flagged for possible exclusion based on focus the study (e.g. modeling tick distributions as an exercise to compare modeling logistics and methodologies), mismatch in target organisms (e.g. the distribution of a target pathogen in a vector of a non-target genus), or status as grey literature (e.g. unpublished theses).

## Results

### Reviewed Studies

The initial search of the peer-reviewed literature yielded 174 studies published between 1994 and 2019. After removal of duplicate search results, 109 unique studies remained. Studies were screened for topic relevance and abstract content, after which 33 studies remained. These publications were then assessed in full, resulting in 24 studies on species distribution models for *Amblyomma* ticks and/or rickettsial pathogens that were included in the literature synthesis (Table 1). The PRISMA flow diagram outlining our literature search and screening process is shown in Fig. 1.

**Table 1.**
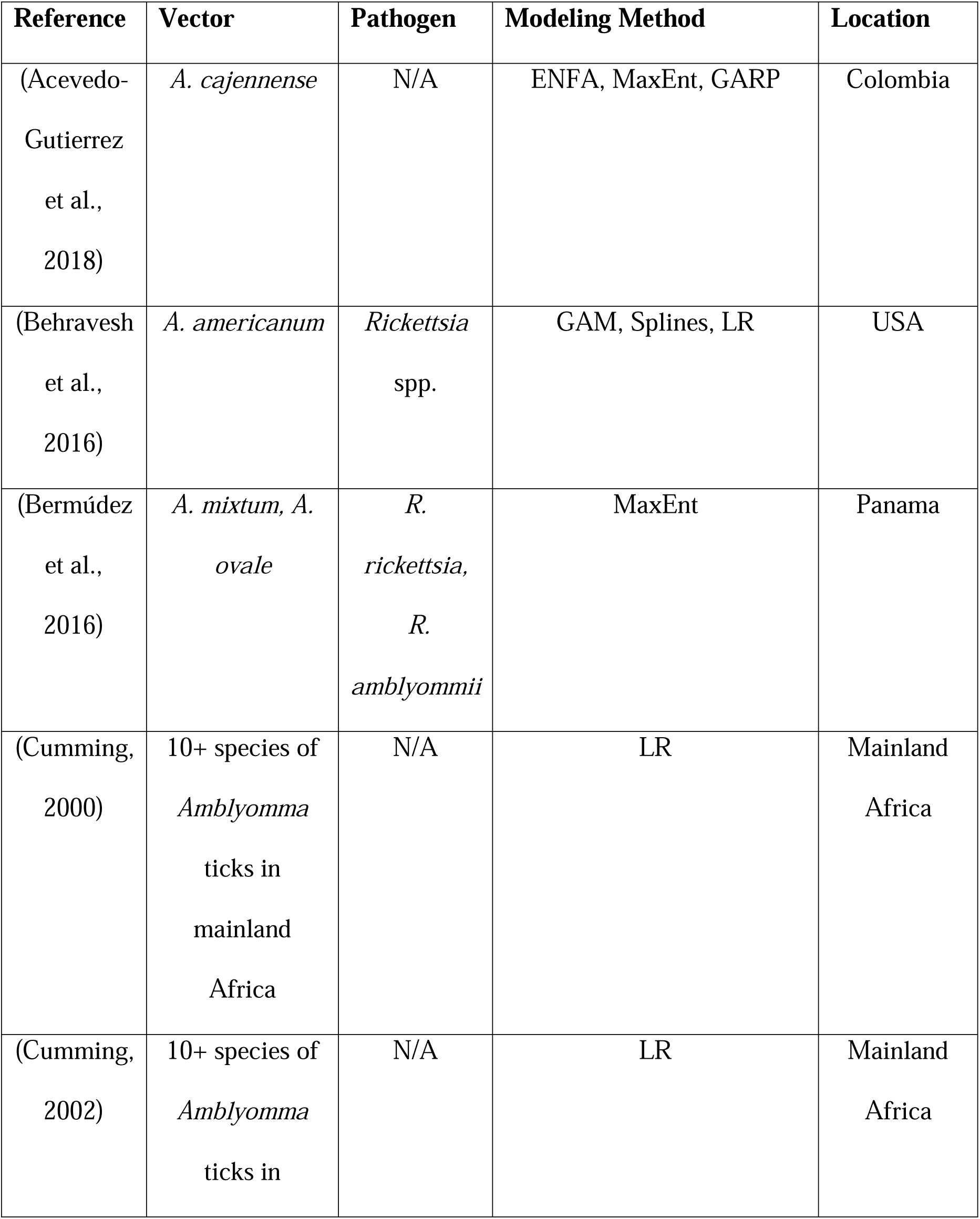
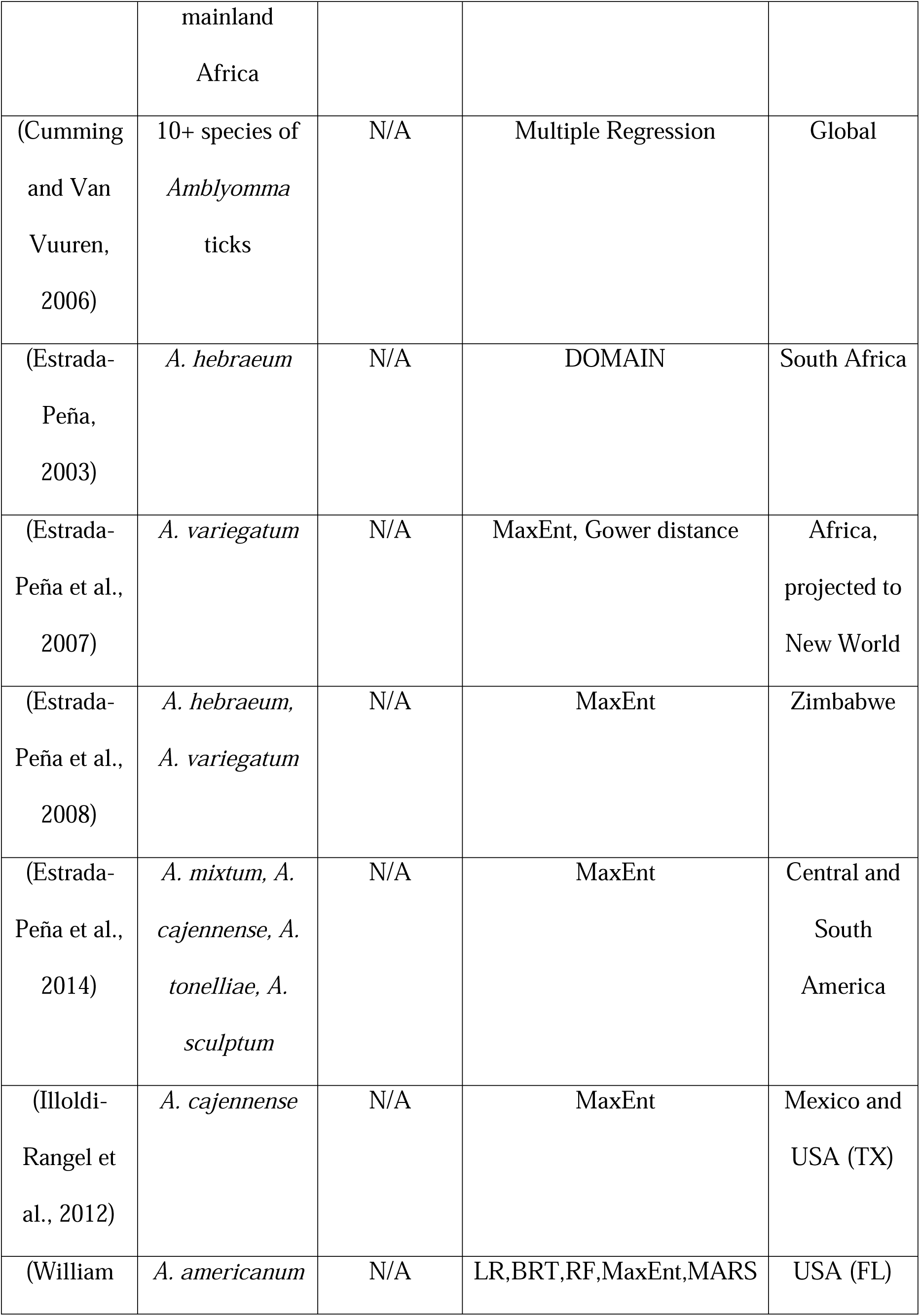
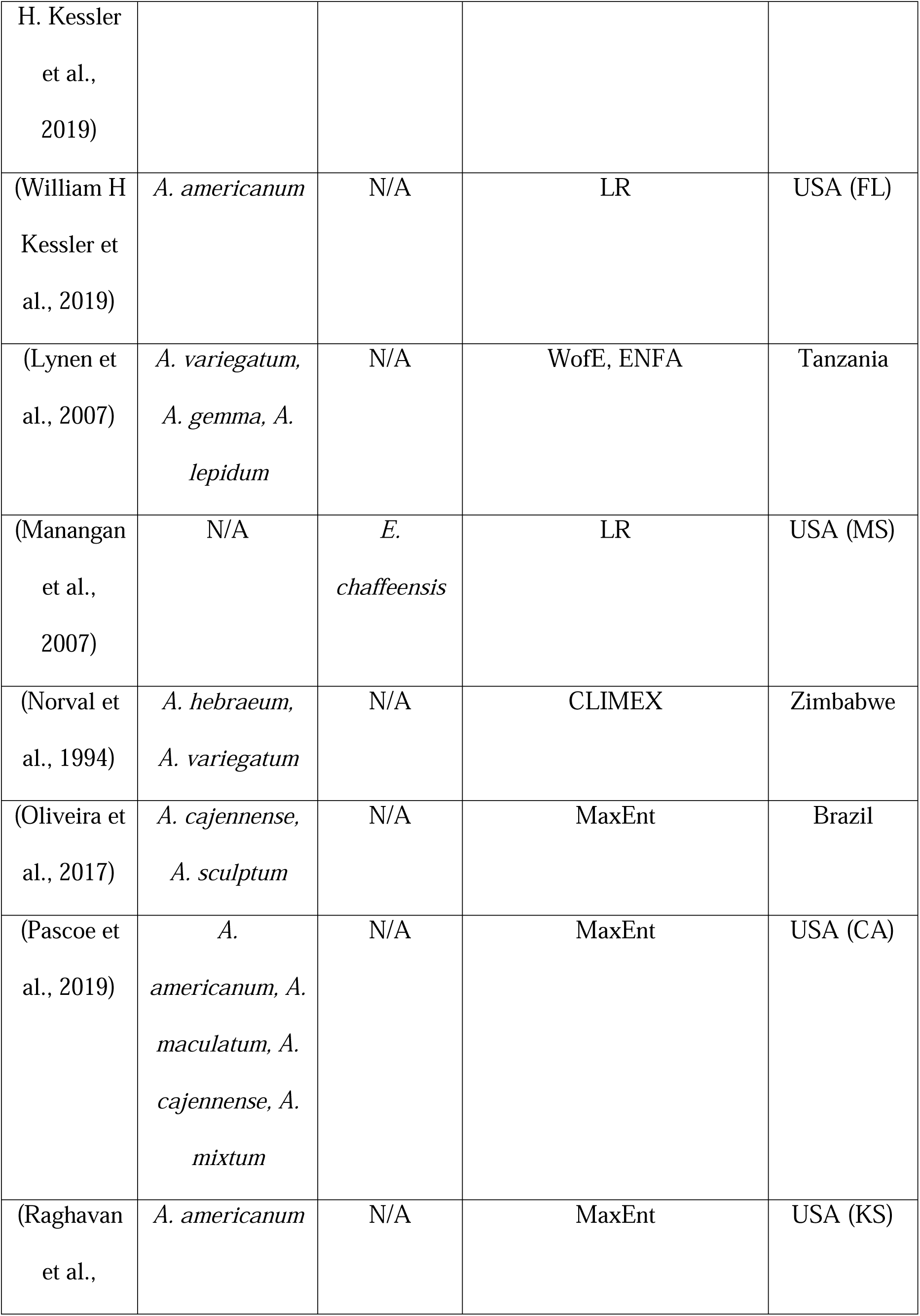
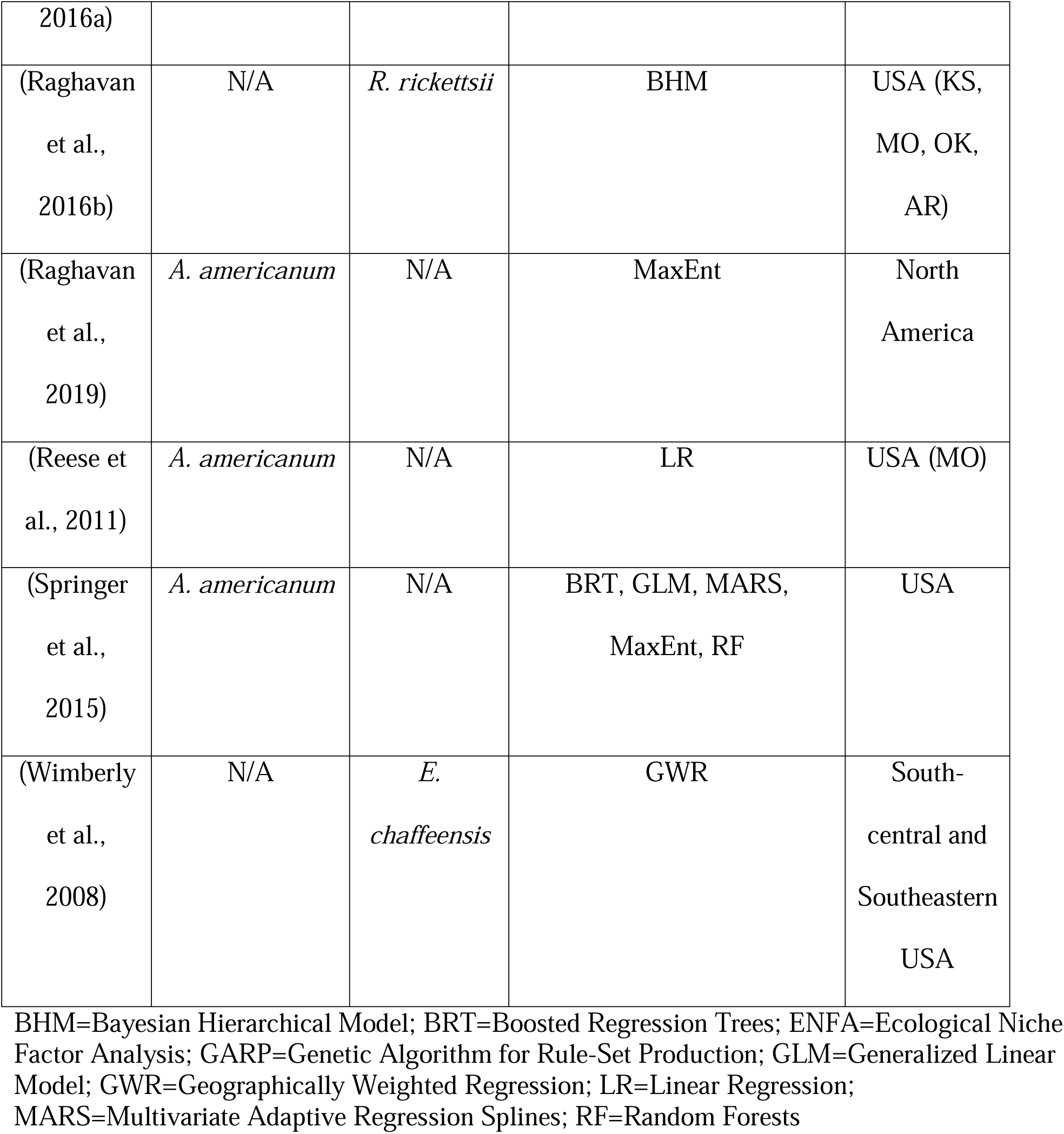
List of final publications on *Amblyomma* ticks and rickettsial group pathogens featured in literature review.

**Figure 1.**
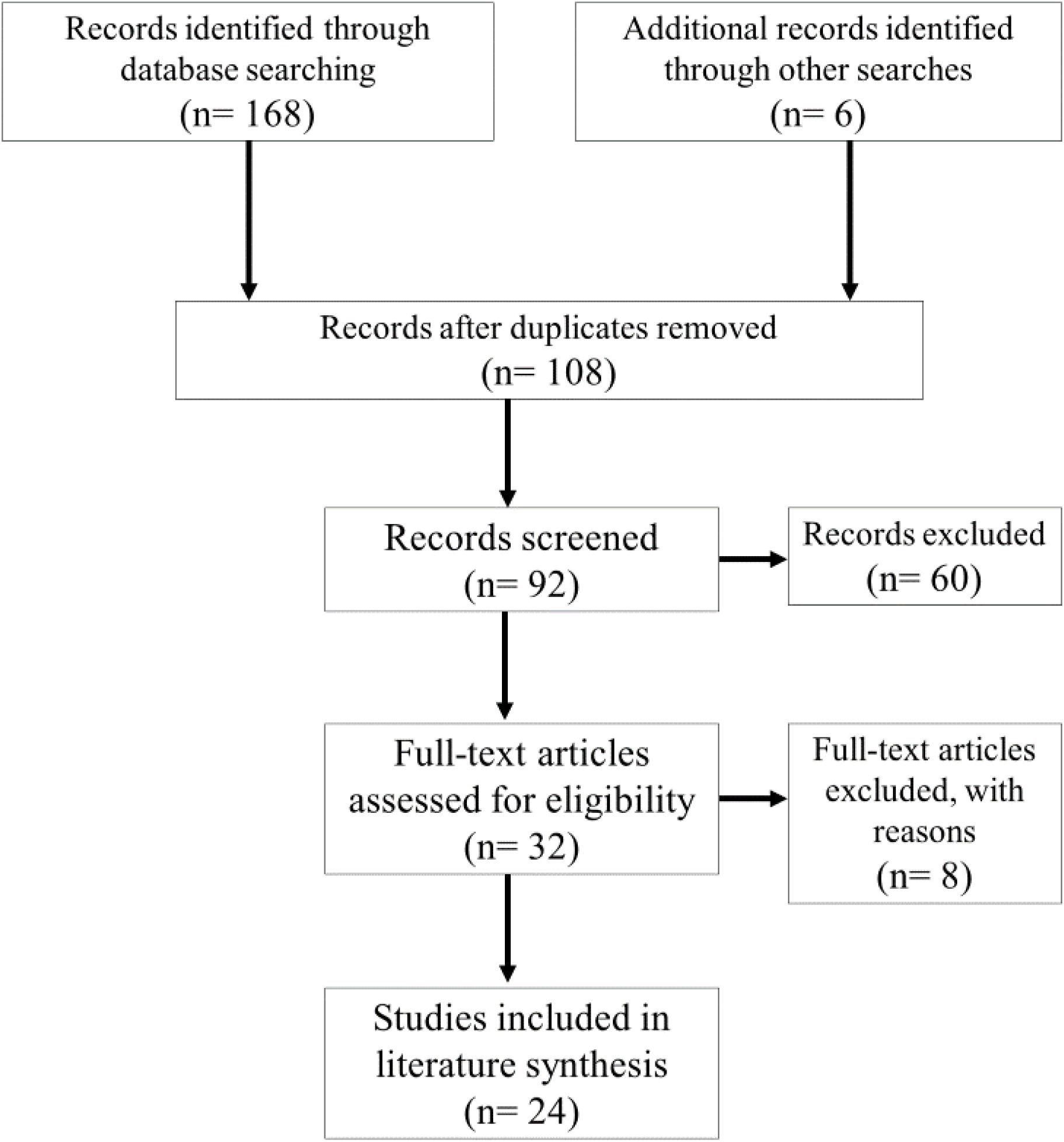
PRISMA flow diagram outlining the literature search and screening process.

The geographic extent of suitability models varied with study focus and stated research goals, ranging from local and regional foci (e.g. county and state-level) to national and global species distributions. Studies were primarily conducted for the United States (46%), often limited to a single state or regional boundary. Other common geographic foci included Africa (29%) and Latin America (21%). The majority of studies (83%) reported the spatial scales of predicted distributions, which varied considerably across studies ranging from fine-scale (1 km) to coarse resolution (50 km) gridded models.

### Species Occurrence Data

The majority of reviewed studies (79%) modeled geographic distributions only for *Amblyomma* ticks, using vector presence as a proxy for pathogen transmission and disease risk. *Amblyomma americanum* was featured in 33% of papers, making it the most commonly studied vector, followed by *Amblyomma variegatum* (29%) and *Amblyomma hebraeum* (25%). Studies that estimated pathogen distributions, using health department data or wildlife blood samples to determine presence, accounted for 20% of the reviewed literature. Only 8% of studies used both tick occurrence and pathogen presence to model geographic risk of transmission.

A majority of studies (63%) obtained positive records of species occurrence from previously published literature. The time period of sample collections from previously published sources typically ranged from the 1950s through the early 2000s, though one study incorporated historical records dating back to the 1900s. In contrast, 29% of studies primarily obtained georeferenced data points from public databases or entomological collections. While using pre-existing databases of tick records yield higher occurrence frequencies that span greater periods of time, few details are typically provided regarding the nature of sample collections (e.g. active versus passive surveillance, transects versus convenience sampling, etc.). Only 21% of studies collected occurrence records solely through field sampling, with field collections spanning one to thirteen years. Few studies (13%) used true absence data collected via field sampling, instead opting to use modeling approaches that take advantage of presence-only datasets.

### Environmental Data

Environmental predictor datasets used to build SDMs were generally chosen in accordance with the specified geographic extent, scale, and goals of a given study. Many of the data products used for localized studies are only available for a given region or country (e.g. Daymet climate data for North America, USGS National Land Cover Database with coverage for the United States, etc.). Despite the wide range of environmental inputs across studies, the WorldClim dataset of long-term climate averages, and derived bioclimatic variables, were the most commonly utilized source of climatological data, featured as input data in 36% of reviewed papers (Hijmans et al., 2005). Six papers estimated potential shifts in vector ranges driven by future climate change and required modeled climate data at given time horizons, with half of these studies using WorldClim scenarios of future climatic conditions.

### Modeling Approaches and Output

The reviewed literature primarily consisted of studies that used presence-only, correlative modeling approaches. A variety of SDM methods were used to estimate tick distributions, including logistic regression (LR), geographically weighted regression (GWR), ecological niche factor analysis (ENFA), and generalized additive models (GAM) (Table 1). However, the MaxEnt algorithm was the most commonly used method, with 50% of studies using MaxEnt to estimate species distributions. Regression analyses were also frequently included in SDM studies (42%), even when more advanced statistical models were also used.

Environmental factors that were most influential in published SDMs were reported in 80% of reviewed studies. Generally, studies included some measure of temperature, precipitation, soil moisture, or land cover in models of tick distributions. The temporal scale of environmental predictors varied considerably between studies, ranging from daily temperature estimates and monthly ranges to annual and long-term climate averages. Covariates that contributed most prominently to distribution estimates were often reported in the reviewed literature, yet actual values and ranges for environmental predictors were seldom reported. Indicators of seasonality for precipitation (66% of studies) and temperature (50% of studies) were among the most consistent covariates included in final SDMs. The Normalized Difference Vegetation Index (NDVI), an index derived from remote sensing data to measure green vegetation cover, was used as an environmental predictor in 25% of studies. Despite the prevalence of NDVI in these studies, compared to climate variables there was little consensus regarding the reliability of this predictor in defining vector niches across studies. Only one study incorporated tick host density as an environmental predictor of habitat suitability.

## Discussion

Species distribution modeling has become a widely used tool for estimating ranges of organisms, or in the case of pathogens and their vectors, the geographic risk of disease exposure. However, the potential geographic distributions of rickettsial pathogens transmitted by *Amblyomma* spp. are still relatively understudied, compared to other vector-borne disease systems. In contrast with the 174 candidate publications identified for screening in this literature review, similar search terminology applied to other vector-borne disease systems yielded raw publication counts of 1,126 for *Aedes* spp. and dengue fever, 728 for *Anopheles* spp. and malaria, and 366 for *Ixodes* spp. and Lyme disease. This lag is likely in part to more recent recognition of the presence of these disease transmission systems for the spotted fever rickettsioses and ehrlichiosis tick-borne diseases (Childs and Paddock, 2003). Correlative models, such as SDMs, are useful in estimating potential species limits, particularly when data for mechanistic or process-driven models are lacking. This is the case for many tick species, where physiological mechanisms and limits are often poorly understood. For *Amblyomma* spp. only a few papers address the effects of temperature and humidity in a controlled setting (Guglielmone, 1992; Koch, 1983). At extreme temperatures where insect vectors may die, ticks will become quiescent until conditions are more favorable. Thresholds that are known to cause instantaneous tick mortality are prohibitive to life such as -22°C (Burks et al., 1996). Presence-only modeling approaches, which predominated in the reviewed literature, are also convenient when studying organisms that are difficult to extensively sample throughout their range. These methods are attractive in that they allow us to take advantage of pre-existing collection datasets, seemingly obviating the need for labor and resource intensive field sampling. However, with the exception of studies which explicitly conducted surveys for ticks, the full methods for originally obtaining presence points (e.g. the original collection strategies) are not always clearly defined. Biases introduced via sampling protocol (e.g. convenience sampling, or targeting a single life stage or behavior) may not adequately represent the true realized niche for species, dramatically influencing SDM predictions. In many tick models, data are collected through dragging or flagging which only samples questing ticks that have not found hosts. The number of successful host-seeking ticks is not known and for ticks that are not possible to collect on drags/flags the surveillance method may drastically underrepresent this life stage (Gaff et al., 2020). If the purpose of the model is to measure disease risk, questing data may be appropriate, but for tick control a greater understanding of the species life history would be needed. While these limitations may be logistically unavoidable, we recommend more detailed reporting of sampling methods and their associated limitations in future studies.

A further difficulty presents itself when estimating species distributions for ticks based on environmental conditions. Other arthropod vectors, such as mosquitoes, are extremely sensitive to fluctuations in climatic conditions, which in turn dictate the suitability of an area for survival and reproduction. In many instances, the physiological responses and environmental limits of insect vectors are well understood, via both laboratory experiments and empirical field studies (Mordecai et al., 2019; Paaijmans et al., 2013; Reuss et al., 2018). In contrast with insect vectors, ticks are resilient to many of the climatic factors that would limit other species. In other words, broad-scale patterns in temperature and precipitation are not necessarily primary drivers of tick presence on the landscape. This may contribute to the relatively low agreement in niche-defining environmental parameters across the reviewed studies. With the exception of indicators of seasonality, the major climatic and land cover predictors in the literature vary greatly with species and geographic extent. Given the close association between ticks and their vertebrate hosts, this may indicate that it is not the vector’s niche that is being modeled, but rather the niche of the host organisms that support tick populations. Furthermore, reaching consensus across SDMs is notoriously problematic, owing largely to the abundance of methodological approaches and lack of standardized reporting practices in presence data and final models (Carlson et al., 2018; Hao et al., 2019; Merow et al., 2013; Mordecai et al., 2019; Rund et al., 2019). We find similar issues when comparing published SDMs for *Amblyomma* ticks and rickettsial pathogens, where there is considerable diversity in methods and primary findings despite the small number of studies performed. While major environmental predictors are typically reported for SDMs, most studies do not report values or numerical ranges for suitability.

## Conclusions

Species distribution modeling of *Amblyomma* ticks and the rickettsial group pathogens they vector is underrepresented in the literature compared to other vector-borne disease systems. Even among a limited number of published studies, there is considerable variation in the methods and reported environmental influences for these models. This systematic literature review highlights a knowledge gap in our understanding of potential geographic risk for this transmission system. Given the recent public health interest in tick-borne diseases, the dearth of studies may result from lags in new data acquisition and limitations in our knowledge of the tick-pathogen ecology involved.

## Data Availability

All data are within the manuscript

## Funding

CAL, HDG, and SJR were funded by NIH 1R01AI136035-01. ALW and SJR were additionally funded by CDC grant 1U01CK000510-01: Southeastern Regional Center of Excellence in Vector-Borne Diseases: The Gateway Program. This publication was supported by the Cooperative Agreement Number above from the Centers for Disease Control and Prevention. Its contents are solely the responsibility of the authors and do not necessarily represent the official views of the Centers for Disease Control and Prevention.

## Abbreviations

SDM: species distribution model
ENM: ecological niche model
PRISMA: Preferred Reporting Items for Systematic Reviews and Meta-analyses
GAM: generalized additive model
BRT: boosted regression trees
RF: random forests
LR: logistic regression (LR)
GWR: geographically weighted regression
ENFA: ecological niche factor analysis

## Notes

### Competing Interest Statement

The authors have declared no competing interest.

